# Exploring Digital Speech Biomarkers of Hypokinetic Dysarthria in a Multilingual Cohort

**DOI:** 10.1101/2022.10.24.22281459

**Authors:** Daniel Kovac, Jiri Mekyska, Vered Aharonson, Pavol Harar, Zoltan Galaz, Steven Rapcsak, Juan Rafael Orozco-Arroyave, Lubos Brabenec, Irena Rektorova

## Abstract

Hypokinetic dysarthria, a motor speech disorder characterized by reduced movement and control in the speech-related muscles, is mostly associated with Parkinson’s disease. Acoustic speech features thus offer the potential for early digital biomarkers to diagnose and monitor the progression of this disease. However, the influence of language on the successful classification of healthy and dysarthric speech remains crucial. This paper explores the analysis of acoustic speech features, both established and newly proposed, in a multilingual context to support the diagnosis of PD. The study aims to identify language-independent and highly discriminative digital speech biomarkers using statistical analysis and machine learning techniques. The study analyzes thirty-three acoustic features extracted from Czech, American, Israeli, Columbian, and Italian PD patients, as well as healthy controls. The analysis employs correlation and statistical tests, descriptive statistics, and the XGBoost classifier. Feature importances and Shapley values are used to provide explanations for the classification results. The study reveals that the most discriminative features, with reduced language dependence, are those measuring the prominence of the second formant, monopitch, and the frequency of pauses during text reading. Classification accuracies range from 67 % to 85 %, depending on the language. This paper introduces the concept of language robustness as a desirable quality in digital speech biomarkers, ensuring consistent behaviour across languages. By leveraging this concept and employing additional metrics, the study proposes several language-independent digital speech biomarkers with high discrimination power for diagnosing PD.

## 1 Introduction

Hypokinetic dysarthria (HD) refers to motor speech disorders manifested in respiration, phonation, articulation, resonance and prosody of speech and has a major impact on the patient’s communication ability [1, 2]. Characteristic features of a person’s voice with HD include tremor [3], hoarseness [4] and breathiness [5]. Speech is further characterised by hypernasality [6], syllable repetitions [7], stuttering [8] and inappropriate silences [9]. Overall, it may be relatively unintelligible [10] and quiet [11], with poor intonation and monoloudness [12]. These disorders most commonly occur in patients with neurodegenerative diseases such as Parkinson’s disease (PD) or various types of dementia. However, stroke, traumatic brain injury, or other conditions that affect the motor control centres in the brain can also lead to the development of HD [13]. In patients with PD, HD is thought to be caused by progressive degeneration of dopaminergic neurons in the substantia nigra [14] and is present in up to 90 % of these people [15]. The risk of PD increases with age and may be influenced by genetic predisposition and various environmental factors [16]. It is also clear that the number of PD patients grows with increasing population and life expectancy [17]. Although the science has made significant advances since the disease was first described [18], we still do not know the actual cause of PD and are not able to cure it; we can only alleviate its symptoms. Therefore, early detection and initiation of treatment are crucial to the future course of the disease [19]. Since HD can begin in the early phases of PD [20], acoustic speech analysis can be a suitable supportive tool for diagnosis or objective monitoring of the disease. Although many teams have worked on this topic, there is still no comprehensive and robust set of acoustic features quantifying the speech disorders that can be applied in practice and capture and describe HD in all domains. One factor that significantly affects these features is the speaker’s language.

In 2010, Whitehill TL [21] described that Chinese PD patients show many of the same patterns of speech abnormalities as English-speaking people with HD and suggested that there may be universal acoustic features that could distinguish between healthy and HD-affected speech.

Hazan et al. (2012) [22] then published the results of a multilingual study focused on the automatic diagnosis of PD patients speaking in English and German. They chose articulatory features based on the formants to distinguish dysarthric from healthy speech. Using the support vector machine as the machine learning model, they were able to predict PD with an accuracy of 85 %, which dropped to 75 % when they trained the model by the features of one language and tested it on the other one. In the summary of the article, they mention that features that can differentiate dysarthric speech from healthy speech probably vary with language.

In 2016, Orozco-Arroyave et al. [23] performed multilingual experiments with speech recordings of Spanish, German, and Czech speakers. The prediction accuracy ranged from 60 % to 99 %, depending on the language combination and the ratio of training to test data. When predicting PD in one language group only, the accuracy ranged from 85 % to 99 %. The most discriminatory features were Mel frequency cepstral coefficients (MFCC) and energy in the critical Bark bands (BBE), both extracted from unvoiced segments of the reading text.

Kim and Choi (2017) [24] published the results of their descriptive study in which they describe the differences in acoustic vowel space (AVS) of Korean- and English-speaking PD patients. No differences in articulation rate were observed.

Next, in 2019 Moro-Velazquez et al. [25] reached an accuracy between 85 % and 94 % when classifying PD patients in a multilingual cohort including Castillian Spanish, Colombian Spanish and Czech. It dropped to the range between 72 % and 82 % when cross-corpora validating. For this purpose, they used an approach based on phonemic grouping. The most significant phonemes for detecting HD were plosives and fricatives. Extraction of the features from reading text led to better results than the quantitative analysis of a diadochokinetic task.

Vásquez-Correa et al. (2019) [26] used convolution neural nets and transfer learning strategy to classify PD in Spanish, German and Czech with MFCC and BBE as input independent variables. Accuracy ranged between 70 % and 77 %. More accurate and more balanced in the frame of sensitivity and specificity were models trained by features of more than one language.

Rusz et al. (2021) [27] performed a speech analysis of Czech, English, German, French and Italian speakers in the early phase of PD. From the sustained phonation of vowel [a], diadochokinetic task and monologue, they extracted seven features in total. They observed significant group differences between PD and controls for monopitch, prolonged pauses, and imprecise consonants. According to statistical analysis, there were no differences between language groups in monopitch and length of pauses.

In 2022, Ozbolt et al. [28] analysed machine learning models trained by phonatory features of Spanish, English and Italian PD patients and healthy controls. These features mainly quantify energies in different parts of spectra of the speech signal recorded during the sustained phonation. The same features were differentially important in English, Spanish and Italian models. During the cross-corpora validating, the accuracy was higher when the model was trained by Spanish data and tested on Italian rather than vice versa. Different vowels of sustained phonation were significant for each scenario.

It is evident that the language has a non-negligible effect on the accuracy of classifying speakers into those who are healthy and those affected with HD, but no study has yet looked at multilingual analysis in depth. This work aims to explore language-independent digital speech biomarkers of PD. It seeks to determine which acoustic speech features are important for different languages in terms of classification accuracy and which are sufficiently robust and independent of the speaker’s language.

## 2 Materials and Methods

The diagram in Figure 1 describes the workflow used to determine the robustness of acoustic speech features to language differences and their subsequent discrimination power. The methods are further specified in the following subsections.

**Figure 1:**
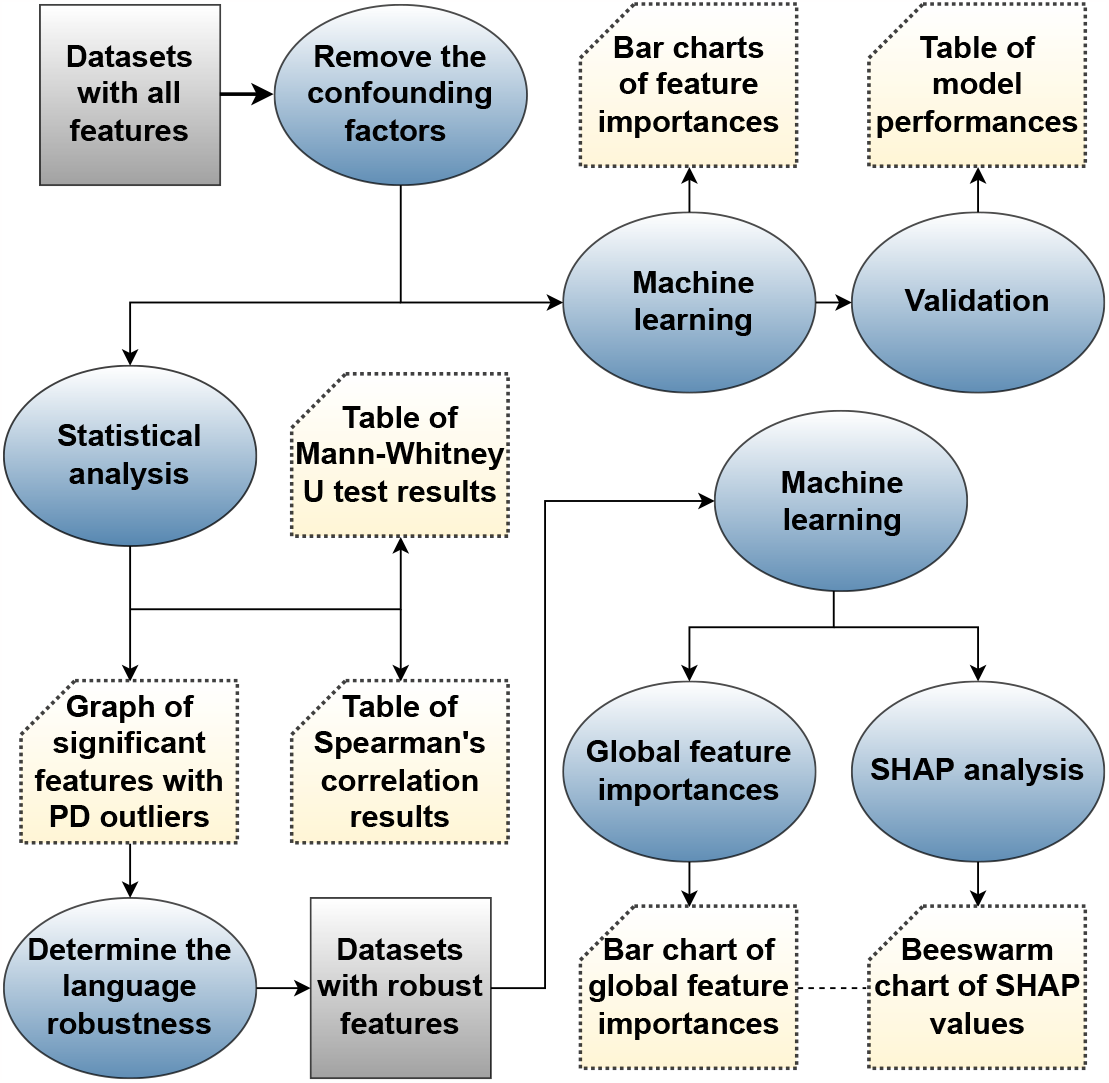
Workflow.

### 2.1 Speech corpus

The corpus contains speech recordings of 506 people (265 healthy controls – HC, 241 PD patients) and is created by grouping several datasets described in Table 1:

**Table 1:** Numbers of PD patients, healthy controls, men (M) and women (F) in the corpus.

|  | CZ |  |  | US |  |  | IL |  |  | CO |  |  | IT |  |  |  |
| --- | --- | --- | --- | --- | --- | --- | --- | --- | --- | --- | --- | --- | --- | --- | --- | --- |
|  | F | M | total | F | M | total | F | M | total | F | M | total | F | M | total | altogether |
| HC | 46 | 35 | 81 | 13 | 5 | 18 | 53 | 43 | 96 | 25 | 25 | 50 | 10 | 10 | 20 | 265 |
| PD | 49 | 84 | 133 | 3 | 8 | 11 | 7 | 12 | 19 | 25 | 25 | 50 | 9 | 19 | 28 | 241 |
| total | 95 | 119 | 214 | 16 | 13 | 29 | 60 | 55 | 115 | 50 | 50 | 100 | 19 | 29 | 48 | 506 |

- Czechs (CZ) including HIDI [29], PARCZ [30] and CoBeN (only HC) [31] data,
- Americans (US) speaking American English from CoBeN project [31],
- Israelis (IL) speaking Hebrew,
- Colombians (CO) speaking Spanish [32],
- Italians (IT) – freely available dataset [33].

Every participant signed informed consent, and the relevant ethics committee approved the study. Their age distribution can be seen in Figure 2 and clinical data of PD patients are summarized in Table 2. This table describes the time since the first symptoms occurred, medication and severity of PD assessed on the Unified Parkinson’s Disease Rating Scale, part III (UPDRS III) and Hoehn and Yahr speech rating scale (H&Y). All patients in the cohort are on dopaminergic medication (ON state). Since deep brain stimulation may affect a person’s speech [34], recordings of patients with this stimulation were discarded from the Israeli dataset, resulting in a slight imbalance of classes that can also be observed in the US data. Two Italian recordings containing a buzzing noise were also removed from the corpus. We analyzed the speech and voice of subjects recorded during a reading of a short text (Read), prolonged phonation of vowel [a], and a diadochokinetic task (DDK) consisting of repeating the syllables [pa]-[ta]-[ka]. The reading task in each dataset contains a different text written in the nation’s corresponding language, and there are two different texts to read in the Czech dataset. In the Italian dataset, patients and healthy controls repeat only the syllable [pa] or the syllable [ta] as a second task variant. In the Czech and American acquisitions, participants performed all tasks only once. In the Israeli dataset, participants had two trials of vowel [a], and in the Colombian one, three trials. The Italians had each task recorded twice. We resampled all recordings to a uniform 16 kHz sampling frequency.

**Table 2:** Clinical data: mean and standard deviation of LED – L-dopa equivalent daily dose; UPDRS III – Unified Parkinson’s Disease Rating Scale, part III (motor examination) and H&Y – Hoehn and Yahr speech rating scale.

|  | CZ | US | IL | CO | IT |
| --- | --- | --- | --- | --- | --- |
| PD duration [year] | $12.2 \pm 5.1$ | $5.0 \pm 2.1$ | $14.1 \pm 6.9$ | $11.2 \pm 9.8$ | U |
| LED [mg] | $1001 \pm 524$ | $286 \pm 222$ | U | U | U |
| UPDRS III | $22.5 \pm 11.5$ | U | U | $37.6 \pm 18.1$ | U |
| UPDRS III speech | U | U | U | $1.3 \pm 0.8$ | $1.0 \pm 1.2$ |
| H&Y | U | U | $2.8 \pm 0.9$ | $2.2 \pm 0.7$ | U |

U – unknown value

**Figure 2:**
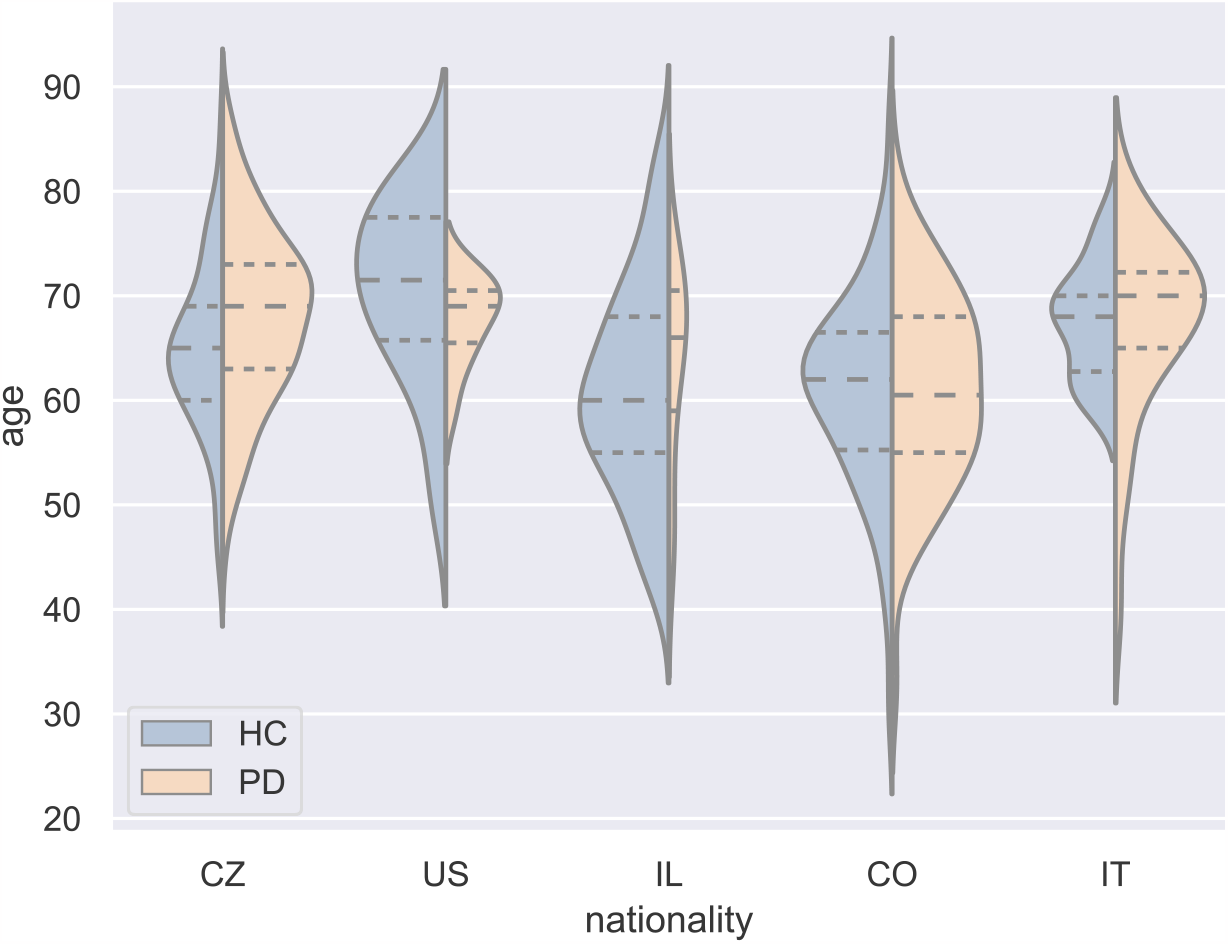
Probability distribution of age.

### 2.2 Parametrization

We extracted 33 acoustic speech features quantifying disorders associated with HD from the recordings. Due to the different signal lengths recorded during vowel [a], we extracted phonatory features from a signal duration of 1.5 s without a vowel onset (keeping a steady state portion). In the case of more than one trial for the speech task, the resulting feature is the average of two or three values obtained from the recordings. Table 3 provides a list and description of these digital biomarkers categorized into phonation (involving respiration), articulation (involving resonance), and prosody (involving timing). This table also describes the expected change in the feature of people with PD. The expectation is based on experience or general knowledge of HD [35, 36, 37]. It is related to the desire to ensure sufficient clinical interpretability, and it also allows us to observe whether a feature behaves as expected in a particular dataset. After parametrization, we regressed out the effect of confounding factors such as age and gender, by training a linear regression model between each confounding factor and a specific acoustic feature. We then used only the residuals of this regression added to the feature mean [38].

**Table 3:**
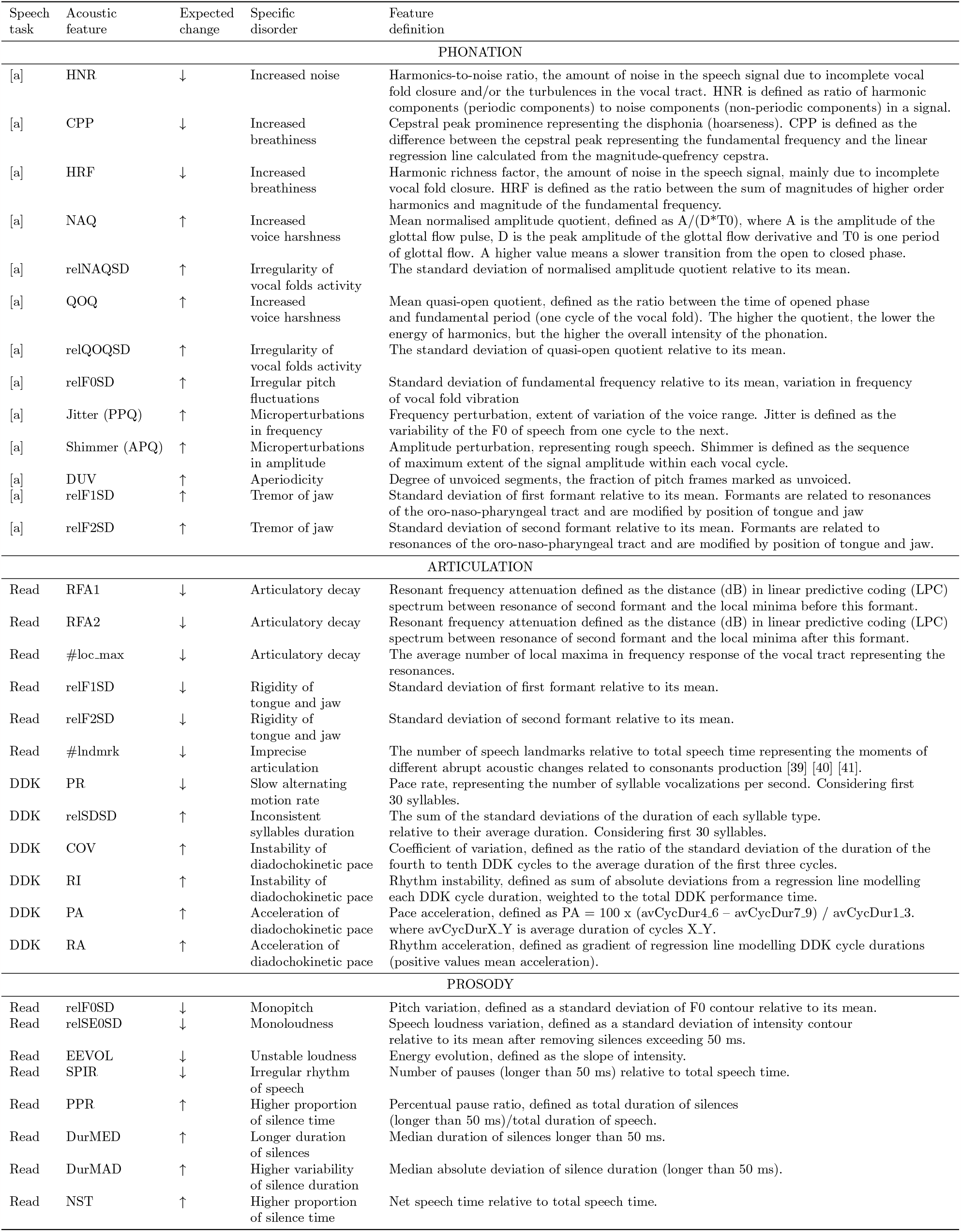
Extracted acoustic features.

### 2.3 Statistical analysis

According to the Shapiro-Wilk test, not all features were normally distributed; therefore, we used the Mann-Whitney U test with the null hypothesis that there is no statistically significant difference in medians of the PD and HC features. Partial Spearman’s rank-correlation coefficients informed us about the negative or positive correlation of the feature with the duration of PD and clinical scores. The language was included as a confounding factor in this test. P-values of all tests were corrected via the False Discovery Rate (FDR) approach. We also looked at the number of patients deviating from the norm given by the distribution of the HC feature. We set the lower *T*_low_ and upper *T*_up_ thresholds to:

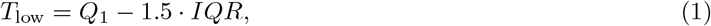

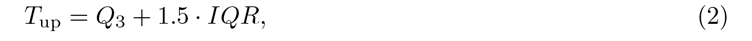

where *Q*_1_ and *Q*_3_ are lower and upper quartiles and *IQR* is the interquartile range. These outliers give us other information about the strength of the feature to detect the HD and show how the feature changes with the disease. Finally, we compared the medians of PD and HC features.

#### 2.3.1 Feature robustness

From the results of the statistical analyses, we determined the language robustness of the features. By language robustness, the exact behaviour of the feature in all datasets is meant. A feature was determined to be robust if it satisfied the following conditions:

1. Significantly discriminated PD from HC in at least one dataset.
2. Followed the assumption in datasets, where it discriminated significantly.
3. The significant correlation of the feature with PD duration and clinical scores went with the assumption.
4. No more than 10 % of PD patients in any dataset deviated from the HC norm against the assumption. The assumption is the expected change in the feature value of people with PD compared to HC (see Table 3 – Expected change).

### 2.4 Machine learning

We chose the Extreme Gradient Boosting (XGBoost) algorithm with a random search hyperparameter tuning strategy to classify speakers into PD patients and HC. We estimated feature importances based on the gain attribute of each feature in the XGBoost model. Before any mathematical modelling, we performed a data transformation first to avoid differences between the features’ values across the language datasets caused by different recording conditions:

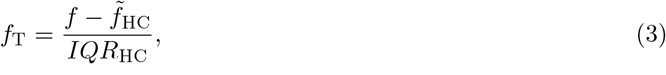

where *f*_T_ is the transformed feature, *f* is an original feature (after the adjustment for age and gender), 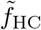_HC_ is a median calculated from HC (in the given language), and *IQR*_HC_ is an interquartile range calculated from HC.

#### 2.4.1 All features

To analyze the effect of the language on classification, we worked with all extracted acoustic features and used three different model validation approaches:

- Stratified 10-fold cross-validation with 20 repetitions
  - 6 scenarios (CZ, US, IL, CO, IT, all)
  stratification ensures balanced train/test data split in the frame of HC/PD and the frame of languages in the scenario with all datasets.
- Cross-language validation technique
  - model trained on data of one dataset and tested on every other.
- Leave-one-language-out validation technique
  - model trained on all but one dataset used for testing.

Models performances were evaluated by Mathews correlation coefficient (MCC), accuracy (ACC), sensitivity (SEN) and specificity (SPE). Feature importances are obtained from models trained on all subjects in each scenario (CZ, US, IL, CO, IT, all), that is, without any split.

#### 2.4.2 Robust features

In the next stage, we took only robust features according to statistical analysis (see section 2.3.1) and trained the models again (CZ, US, IL, CO, IT) to get feature importances. Multiplying the importance coefficients of each model with subsequent normalisation gives the global importance coefficients that we ranked in order to find the most robust features. Finally, we trained the model on all datasets and analysed it with the SHAP approach based on game theory to observe the feature behaviour when classifying subjects speaking in different languages.

## 3 Results

### 3.1 Statistical analysis

Table 4 describes the results of the Mann-Whitney U test and the change in statistical parameters of PD patients compared to HC. Highlighted are the p-values of the features that significantly differentiate these classes. Table 5 shows the results of the correlation of each feature with the duration of PD, medication and clinical scores. Highlighted p-values represent significant correlations. The significance level for rejecting the null hypothesis was 0.05 for both tests. Figure 3 summarizes the features that exhibit statistically significant differences between the classes’ medians, as determined by the Mann-Whitney U test, and assesses the features’ robustness across languages.

**Table 4:**
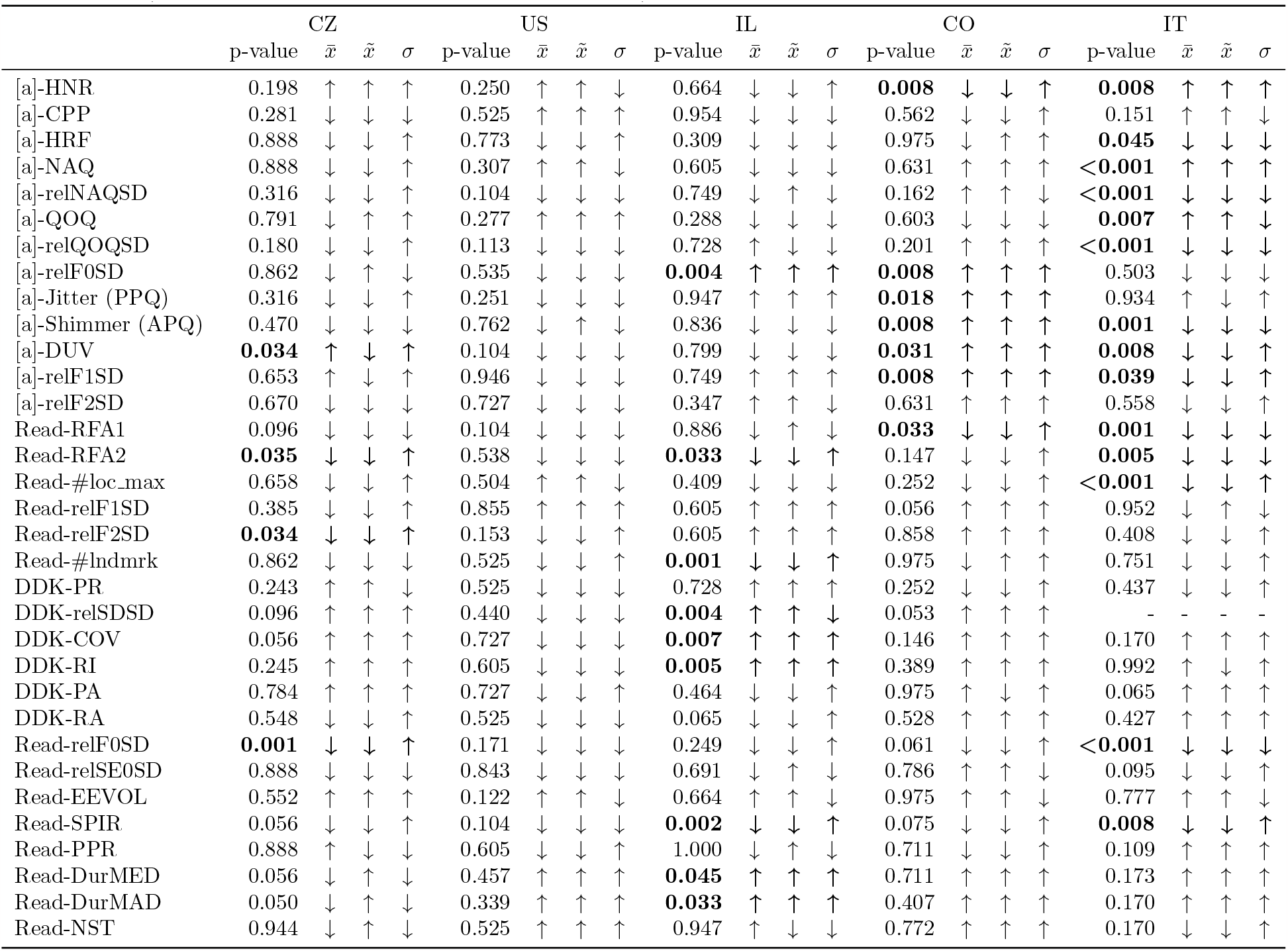
Results of Mann-Whitney U test (p-values after the FDR correction) and changes in statistical parameters (mean 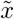, median 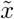, standard deviation *σ*) with PD.

**Table 5:** Results of partial Spearman’s rank correlation (p-values after the FDR correction).

|  | PD duration |  | LED |  | UPDRS III |  | UPDRS III speech |  | H&Y |  |
| --- | --- | --- | --- | --- | --- | --- | --- | --- | --- | --- |
|  | coeff | p-value | coeff | p-value | coeff | p-value | coeff | p-value | coeff | p-value |
| [a]-HNR | -0.192 | **0.032** | 0.161 | 0.347 | -0.081 | 0.471 | -0.090 | 0.556 | -0.065 | 0.860 |
| [a]-CPP | -0.189 | **0.030** | 0.089 | 0.610 | -0.038 | 0.774 | -0.015 | 0.957 | -0.044 | 0.982 |
| [a]-HRF | -0.071 | 0.442 | -0.052 | 0.775 | -0.244 | **0.016** | -0.100 | 0.550 | -0.109 | 0.778 |
| [a]-NAQ | 0.067 | 0.444 | 0.092 | 0.610 | 0.217 | **0.037** | 0.235 | 0.113 | 0.119 | 0.778 |
| [a]-relNAQSD | 0.236 | **0.009** | -0.157 | 0.347 | 0.033 | 0.774 | 0.017 | 0.957 | 0.007 | 0.982 |
| [a]-QOQ | -0.041 | 0.669 | -0.071 | 0.660 | 0.022 | 0.857 | -0.029 | 0.916 | -0.015 | 0.982 |
| [a]-relQOQSD | 0.255 | **0.005** | -0.166 | 0.347 | 0.014 | 0.865 | -0.007 | 0.957 | 0.049 | 0.954 |
| [a]-relF0SD | -0.079 | 0.395 | 0.103 | 0.609 | 0.013 | 0.865 | 0.098 | 0.550 | -0.021 | 0.982 |
| [a]-Jitter (PPQ) | 0.153 | 0.094 | -0.023 | 0.945 | 0.053 | 0.631 | 0.239 | 0.113 | 0.014 | 0.982 |
| [a]-Shimmer (APQ) | 0.214 | **0.017** | -0.131 | 0.491 | 0.148 | 0.154 | 0.104 | 0.550 | 0.084 | 0.857 |
| [a]-DUV | 0.090 | 0.395 | -0.009 | 0.952 | 0.042 | 0.726 | 0.148 | 0.391 | -0.068 | 0.860 |
| [a]-relF1SD | 0.196 | **0.030** | -0.185 | 0.347 | 0.060 | 0.583 | 0.418 | **0.003** | 0.000 | 0.999 |
| [a]-relF2SD | -0.146 | 0.099 | 0.005 | 0.952 | 0.013 | 0.865 | 0.182 | 0.256 | 0.024 | 0.982 |
| Read-RFA1 | 0.007 | 0.921 | -0.059 | 0.768 | -0.190 | 0.057 | -0.085 | 0.569 | -0.113 | 0.778 |
| Read-RFA2 | -0.283 | **0.002** | 0.089 | 0.610 | -0.154 | 0.140 | -0.071 | 0.640 | -0.103 | 0.780 |
| Read-#loc_max | -0.184 | **0.039** | 0.100 | 0.609 | -0.114 | 0.295 | -0.144 | 0.391 | -0.160 | 0.705 |
| Read-relF1SD | 0.035 | 0.684 | -0.055 | 0.775 | 0.021 | 0.857 | -0.121 | 0.517 | 0.030 | 0.982 |
| Read-relF2SD | -0.142 | 0.109 | 0.014 | 0.952 | -0.080 | 0.471 | -0.148 | 0.391 | 0.009 | 0.982 |
| Read-#lndmrk | -0.213 | **0.017** | -0.007 | 0.952 | -0.113 | 0.295 | -0.353 | **0.015** | -0.175 | 0.705 |
| DDK-PR | 0.134 | 0.135 | 0.037 | 0.877 | 0.204 | **0.039** | 0.093 | 0.556 | 0.130 | 0.778 |
| DDK-relSDSD | 0.171 | 0.059 | 0.117 | 0.501 | 0.265 | **0.011** | 0.323 | 0.077 | 0.305 | 0.215 |
| DDK-COV | 0.148 | 0.137 | 0.145 | 0.491 | 0.216 | 0.063 | 0.394 | **0.019** | 0.306 | 0.215 |
| DDK-RI | 0.025 | 0.779 | 0.073 | 0.660 | 0.156 | 0.140 | 0.185 | 0.256 | 0.130 | 0.778 |
| DDK-PA | -0.054 | 0.684 | 0.132 | 0.630 | 0.103 | 0.579 | 0.349 | 0.256 | 0.102 | 0.860 |
| DDK-RA | 0.068 | 0.444 | -0.123 | 0.491 | -0.102 | 0.354 | -0.006 | 0.957 | -0.013 | 0.982 |
| Read-relF0SD | -0.018 | 0.826 | -0.009 | 0.952 | -0.211 | **0.037** | -0.100 | 0.550 | 0.078 | 0.860 |
| Read-relSE0SD | 0.149 | 0.098 | 0.073 | 0.660 | 0.112 | 0.295 | 0.108 | 0.550 | -0.153 | 0.705 |
| Read-EEVOL | 0.078 | 0.395 | 0.284 | **0.022** | 0.033 | 0.774 | 0.012 | 0.957 | -0.163 | 0.705 |
| Read-SPIR | -0.065 | 0.445 | 0.049 | 0.776 | -0.093 | 0.414 | -0.530 | **<0.001** | -0.278 | 0.215 |
| Read-PPR | 0.085 | 0.395 | 0.124 | 0.491 | 0.090 | 0.419 | 0.290 | **0.048** | 0.109 | 0.778 |
| Read-DurMED | 0.078 | 0.395 | 0.021 | 0.945 | 0.121 | 0.289 | 0.353 | **0.015** | 0.243 | 0.303 |
| Read-DurMAD | 0.084 | 0.395 | -0.028 | 0.945 | 0.122 | 0.289 | 0.335 | **0.019** | 0.280 | 0.215 |
| Read-NST | -0.080 | 0.395 | -0.091 | 0.610 | -0.065 | 0.579 | -0.285 | **0.048** | -0.096 | 0.800 |

**Figure 3:**
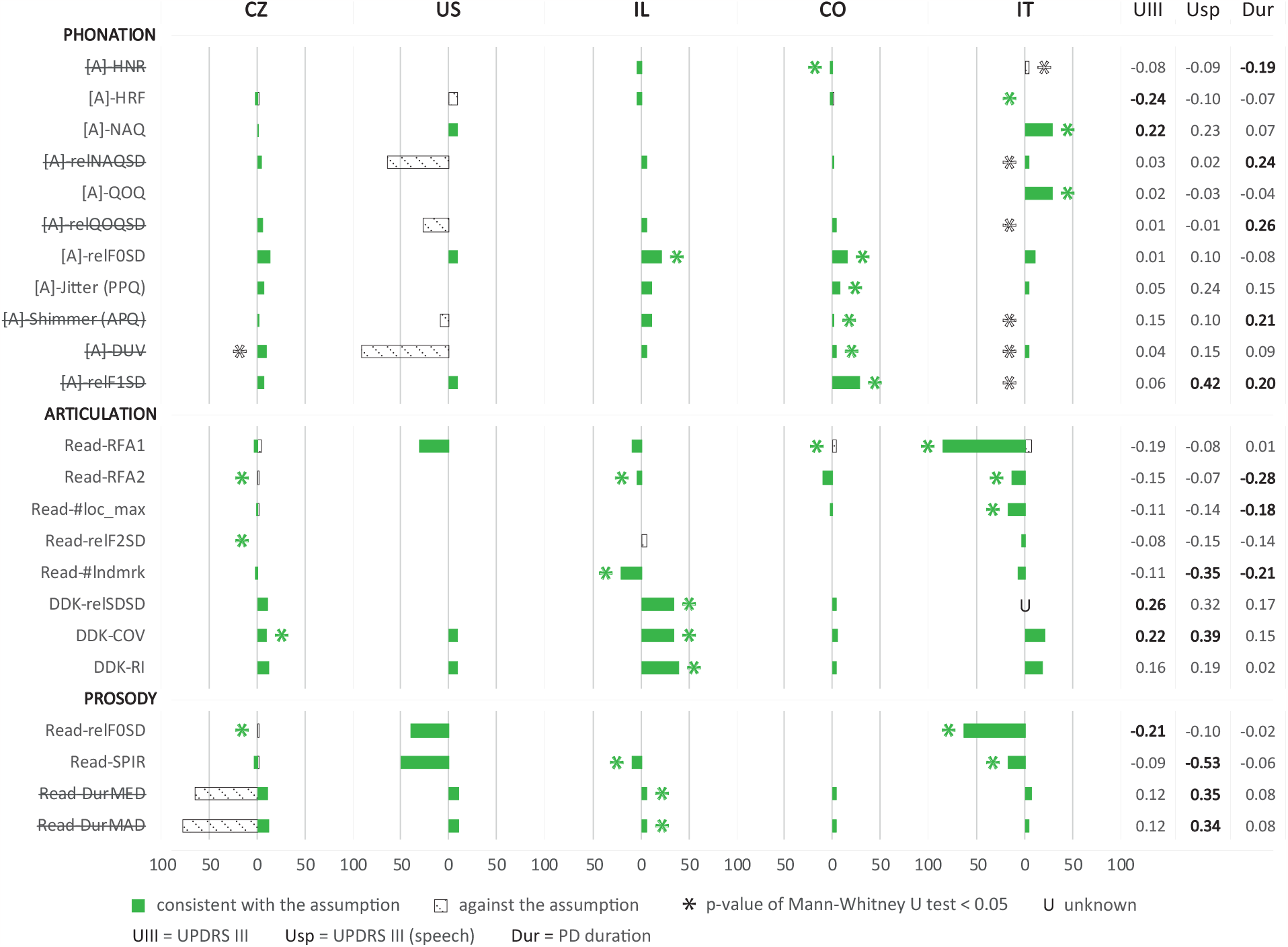
Percentage ratio of PD patients outside the norms given by HC. The left side from zero shows the percentage of PD patients below the lower limit and the right side above the upper limit. The significant difference between HC and PD represents an asterisk, and its position indicates the direction in which the median value of the PD feature has changed compared to HC. Spearman correlation coefficient values are highlighted if they are statistically significant (p-value *<* 0.05). Features not following the condition for being robust are crossed out.

### 3.2 Machine learning

The performance of each machine learning model trained by all features can be seen in Table 6: 10-fold cross-validation with 20 repetitions, Table 7: cross-language validation and Table 8: leave-one-language-out. In Figure 4, we ranked the acoustic features according to their importance in different language scenarios, and Figure 5 shows the global importance of the robust features only and their behaviour in the model trained by the robust features of all subjects in the corpus.

**Table 6:** Classification results: stratified cross-validation.

|  | MCC | ACC [%] | SEN [%] | SPE [%] |
| --- | --- | --- | --- | --- |
| CZ | $0.28 \pm 0.21$ | $67 \pm 9$ | $82 \pm 11$ | $43 \pm 18$ |
| US | $0.42 \pm 0.54$ | $70 \pm 26$ | $74 \pm 42$ | $68 \pm 37$ |
| IL | $0.37 \pm 0.35$ | $79 \pm 11$ | $58 \pm 36$ | $83 \pm 11$ |
| CO | $0.43 \pm 0.27$ | $70 \pm 13$ | $62 \pm 21$ | $78 \pm 20$ |
| IT | $0.71 \pm 0.31$ | $85 \pm 5$ | $86 \pm 19$ | $83 \pm 27$ |
| all | $0.49 \pm 0.11$ | $75 \pm 6$ | $73 \pm 8$ | $76 \pm 7$ |

**Table 7:**
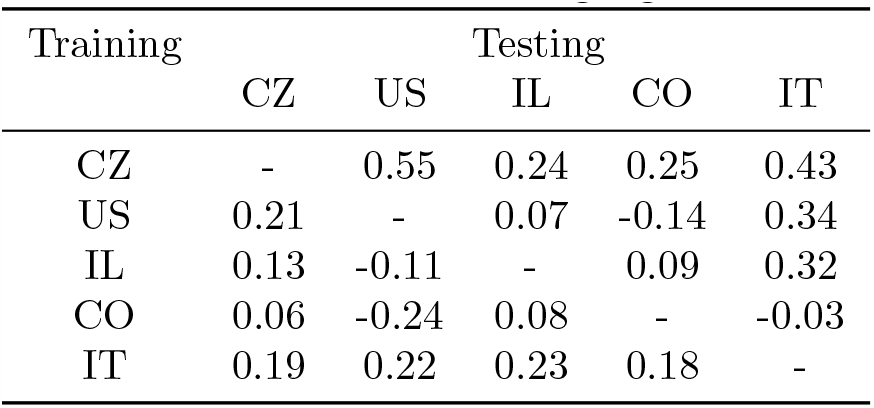
Classification results: cross-language validation – MCC.

**Table 8:** Classification results: leave-one-language-out.

| Testing | MCC | ACC [%] | SEN [%] | SPE [%] |
| --- | --- | --- | --- | --- |
| CZ | 0.19 | 58 | 53 | 67 |
| US | 0.48 | 76 | 45 | 94 |
| IL | 0.30 | 53 | 95 | 45 |
| CO | 0.00 | 50 | 78 | 22 |
| IT | 0.61 | 79 | 71 | 90 |

**Figure 4:**
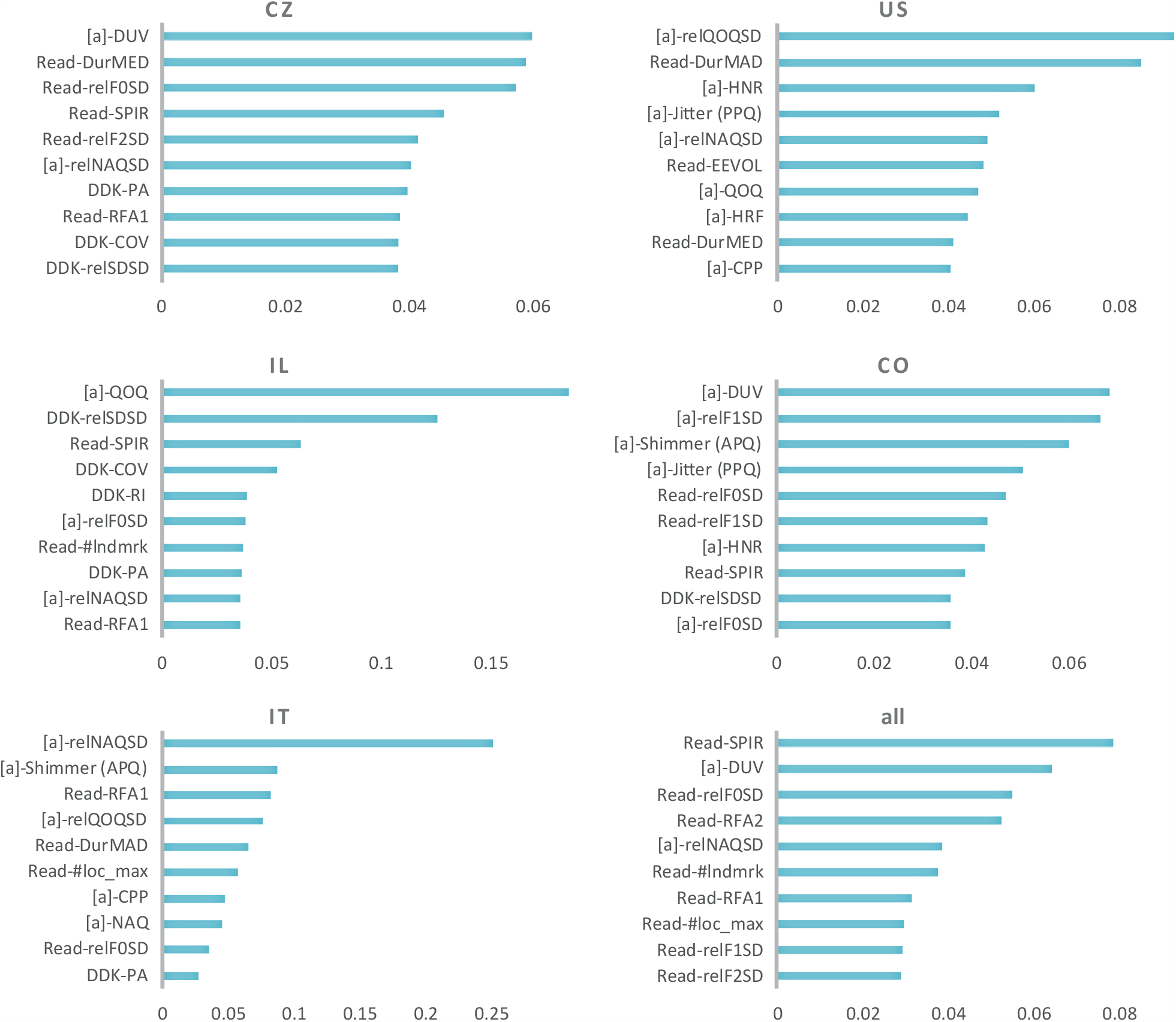
Coefficients of importance of ten most important features in each language scenario (machine learning models trained by all extracted features).

**Figure 5:**
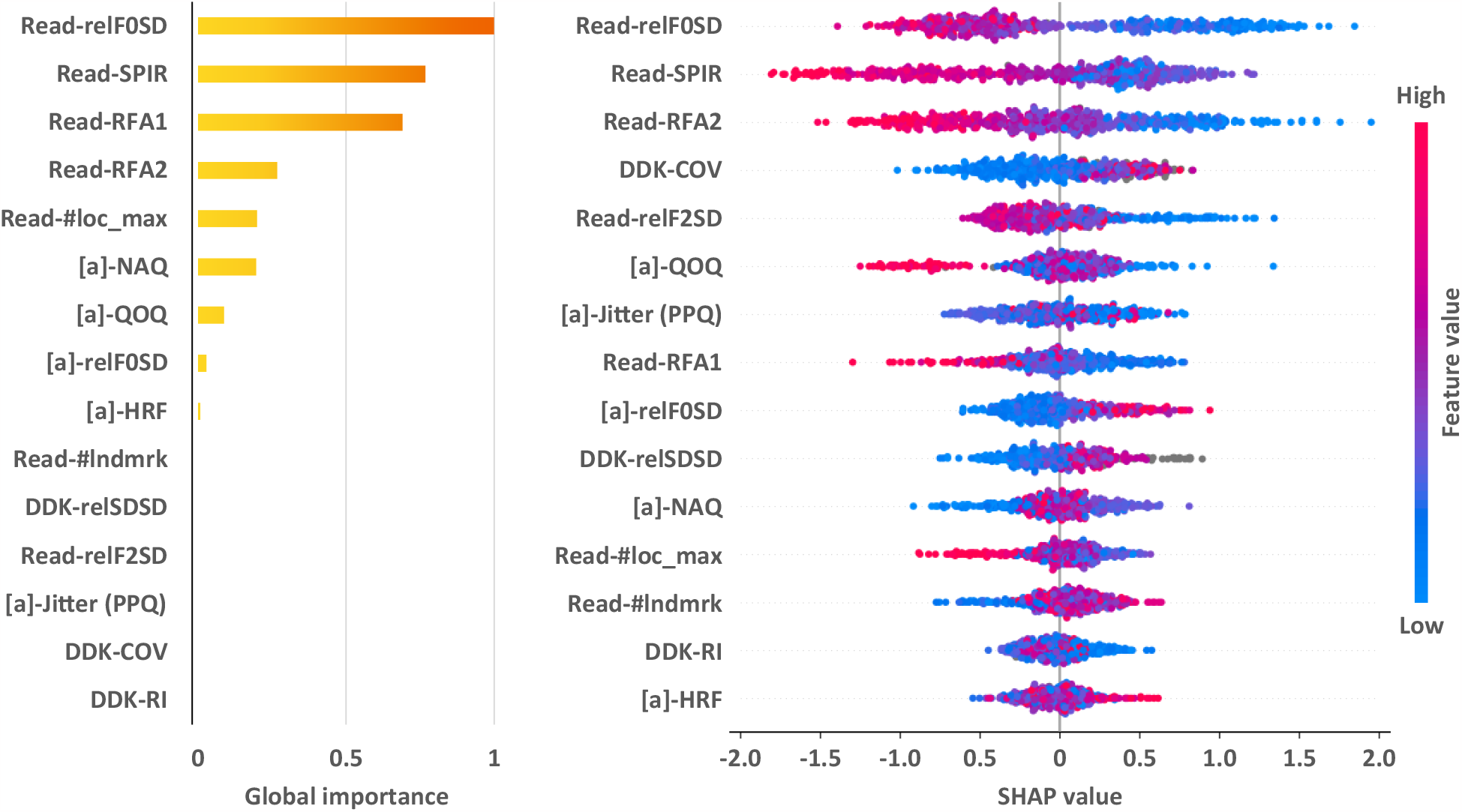
Global normalized combination of importance coefficients of robust features **(left)** and description of the model trained by robust features of all languages together using SHAP values **(right)** (features ordered from the highest absolute SHAP value, negative values indicate a higher probability for the classification to HC and positive values to patients with PD).

## 4 Discussion

We processed voice and speech recordings of 506 individuals (265 HC, 241 PD) speaking five different languages to identify language-independent speech biomarkers with high discriminative power. The corpus includes Czech (CZ), American English (US), Hebrew (IL), Colombian Spanish (CO) and Italian (IT). Acoustic features, quantifying phonatory, articulatory and prosodic disorders, were extracted from the signal recorded during text reading (Read), prolonged phonation of vowel [a] and diadochokinetic task (DDK). We then performed a statistical analysis to obtain the language robustness of the features, followed by machine learning to observe the impact of language on classification accuracy, focusing on individual speech features. We compared the Mathews correlation coefficient (MCC), accuracy (ACC), sensitivity (SEN) and specificity (SPE) of specific models.

### 4.1 Statistical analysis

According to the Mann-Whitney U test, most features discriminate significantly in the Italian dataset (14/33), following the Israeli (9/33) and Spanish (7/33) ones. There are four significant features in the Czech dataset and none in the American one. Of the 33 extracted acoustic speech features, 23 discriminated significantly in at least one language dataset (Table 4), after which seven did not meet the language robustness conditions (Figure 3).

Despite the expected deterioration of speech in PD patients, some features indicated better performance in some language groups. In the Italian dataset, compared to healthy controls, patients had a significantly less breathy voice (based on the feature [a]-HNR) with fewer irregularities ([a]-relNAQSD, [a]-relQOQSD) and fewer perturbations in amplitude ([a]-Shimmer (APQ)). At the same time, they exhibited reduced tremor of jaw ([a]-relF1SD), and their speech was more voiced ([a]-DUV). A significantly higher proportion of voiced segments in patients also occurred in the CZ dataset. Moreover, in the US dataset, many patients deviated from the norms of healthy controls against the assumption in the feature [a]-relNAQSD (64 %), [a]-relQOQSD (27 %) and [a]-DUV (91 %). For these reasons, the mentioned phonatory features did not meet the conditions for sufficient language robustness. We also enclosed prosodic features quantifying the duration of pauses (Read-DurMED) and the variation of their duration (Read-DurMAD) due to the high number of patients deviating from the HC norms against the assumption (64 % and 79 %) in the CZ dataset. However, this dataset comprises several sub-datasets, one of which involves people reading different text compared to the other two sub-datasets. Assuming a non-equal distribution of HC/PD subjects across these three sub-datasets, we attribute the deviation to this factor.

Most features did not meet the robustness conditions due to the Italian dataset, where PD patients performed better than HC in half of the phonatory features. The Italians might manifest HD differently in their voice and breathing. Another explanation can be that the phonation of these patients is positively affected by medication, and they may belong to a specific subtype of HD, as described in the study by Rusz et al. [42]. In this study they also present monopitch as a feature that is common in each subtype and because our results show language robustness in this feature, it gains a high potential in the field of objective HD assessment. Moreover, in another of their studies [27], this feature was also independent of the speaker’s language based on the general least-squares linear models and had significant discrimination power. Other aspects of PD clinical heterogeneity have to be taken into consideration, such as tremor dominant vs hypokinesia/rigidity/gait instability dominant subtypes.

The successful significant and language-independent biomarkers on the basis of statistical analyses hence remain the following features: increased breathiness due to incomplete vocal fold closure ([a]-HRF), increased voice harshness due to a slower transition from an open to a closed phase ([a]-NAQ) and a longer duration of an open phase ([a]-QOQ) of the vocal fold cycle, irregular pitch fluctuations ([a]-relF0SD), microperturbations in frequency ([a]-Jitter (PPQ)), articulatory decay as lower prominence of the second formant (Read-RFA1, Read-RFA2) and less local maxima (Read-#loc max) in the LPC spectrum, the rigidity of tongue and jaw as the lower standard deviation of the second formant during the reading (Read-relF2SD), imprecise articulation due to a lower number of speech landmarks (Read-#lndmrk), inconsistent syllables duration as a higher variance of syllable duration during the diadochokinetic task (DDK-relSDSD), instability of diadochokinetic pace as an increased variance of the cycle duration at the end of the task compared to the beginning (DDK-COV) and overall variance of cycle duration (DDK-RI), monopitch (Read-relF0SD) and a lower number of pauses during the speech (Read-SPIR). These are the features that proved consistent behaviour across all language datasets.

There was no significant correlation of any feature with the H&Y score (Table 5), and only one feature (Read-EEVOL) correlated significantly with medication. A positive Spearman’s coefficient indicates better loudness stability in PD patients with higher LED. However, more features are correlated with the remaining clinical data, such as PD duration, UPDRS III, and UPDRS III speech. The feature Read-RFA2 (articulatory decay), which is the only one that demonstrated significant discrimination across three datasets (CZ, IL, and IT), exhibited a strong correlation with the disease duration, yielding the most promising results.

Looking at the results, it is evident that the ability of acoustic speech features to detect HD is indeed language-dependent [22]. However, it is worth noting that certain features manifest similarly with the development of HD in different language groups. This observation supports the original hypothesis proposed by Whitehill [21], suggesting the existence of a language-universal component of dysarthria.

### 4.2 Machine learning

Based on all extracted features, we were able to detect PD with accuracy ranging from 67 % to 85 %, depending on the language dataset. For all languages combined, the model achieved an accuracy of 75 % with a sensitivity of 73 % and a specificity of 76 % (Table 6). The accuracy here is approximately the average of the values of each language model. With lower standard deviations of cross-validation results and higher balance in terms of sensitivity and specificity, a classifier based on gradient trees can be considered suitable when dealing with multilingual data. Vasquez-Correa et al. [26] also observed a lower difference between sensitivity and specificity and lower standard deviation of cross-validation results when fine-tuning the deep machine learning model. This implies that the model trained on more data has a more balanced classification despite the language differences when choosing an appropriate machine learning approach. However, to maintain high classification accuracy, it is probably necessary that the subjects that the model classifies are from the language group that was included during the training process. Otherwise, the classification results decrease rapidly. We can observe this phenomenon in dropped MCC during cross-language validation (Table 7). The only improvement was an increase in MCC values from 0.42 to 0.55 in the classification of the US subjects when the model was trained on CZ data. Although the small size of the US dataset needs to be considered here, the value of MCC dropped from 0.42 to -0.24 when the US was classified by the model trained on CO data. It supports the hypothesis of the dependence of successful classification of PD on the chosen source and target language group during cross-language validation. This should be taken into account in the possible use of transfer learning. Overall, the classifications were most successful when the model was trained on CZ data and worst when trained on CO data. Next to the combination of CO and any other language, the combination of IT and US seems also inappropriate. The leave-one-language-out validation approach results support the previous findings (Table 8) – classification of US subjects is the only one that shows better results; MCC increased from 0.42 to 0.48, which is a slightly lower improvement compared to the model trained on CZ data only. The drop in accuracy when classifying people speaking a different language to subjects used during training is consistent with previous studies’ results [22, 23, 25], but it appears that data from another carefully-selected language can be used to improve the model performance. From the importance coefficients of each model (Figure 4), we can observe that in the classification, features are differently important for each scenario, which confirms the earlier findings [28]. Counting the occurrences of features in all feature importance scenarios, the best results are provided by the feature [a]-relNAQSD, which is important in the model trained on all languages and in four out of five separate models. However, this feature is not robust according to previous statistical analysis (see Figure 3). The other features that yield the best results in this regard are Read-SPIR, Read-relF0SD, and Read-RFA1. These features all appeared in the feature importances of the model trained on all languages and in three other scenarios. Moreover, all of these features are considered robust based on statistical analysis. After training the machine learning models with only the robust features, globally, the most important features are again monopitch (Read-relF0SD), inappropriate silences (Read-SPIR) and articulatory decay (Read-RFA1) (Figure 5). From the SHAP values of the model trained on data of all languages, it is clear that some features of high importance in the individual models lose their ability to discriminate here (Read-#loc max, [a]-NAQ, [a]-HRF). The features that have been most effective in this model are Read-relF0SD and Read-SPIR once again. In third place, according to the absolute SHAP value, is the articulatory decay (Read-RFA2). Thus, reading text appears to be the crucial speech exercise for successful classification, which is consistent with the results of the study by Moro-Velazquez et al. [25]. Furthermore, speech pausing and rhythmicity abnormalities are associated with cognitive dysfunction in advanced PD stages [43] [44] and can serve as a predictive marker for the cognitive decline in PD patients as well [45]. These abnormalities, along with articulatory muscle bradykinesia with temporal decrements and monopitch, have been previously described by us [34] and others [46] as early markers of HD in PD.

### 4.3 Limitations of the study

This study has several limitations, the most major of which is the heterogeneity of the datasets. Each language dataset has a different number of subjects. There are only 29 subjects in the American dataset, which is 185 subjects less than in the Czech one. The US patients also have the disease for a much shorter time, which can explain why no feature discriminates significantly in this group. Moreover, since some groups’ clinical data (mainly LED, duration of the disease, and dysarthria severity) are unknown, we cannot tell whether subjects in different datasets have the same level of disease progression. The fact that the voice and speech of the subjects in each dataset were recorded using a different acquisition protocol and hardware (e.g., different microphones, microphone-to-mouth distances, etc.) could also play a role, even though we attempted to mitigate this effect by applying feature transformation based on healthy control subjects in the specific language. Due to the different ways of performing speech tasks in each country, we did not test whether individual features from different language groups come from the same probability distribution. The different approach to the diadochokinetic task for the Italian acquisition also caused the inability to extract the feature quantifying the inconsistent syllables duration as the sum of the standard deviations of the duration of each syllable type (DDK-relSDSD). Furthermore, missing the monologue exercise in some datasets made it impossible to use this speech task in our multilingual study. We also need to consider that the classes (especially in the Israeli dataset) are unbalanced, which can mostly, along with the different sizes of datasets, affect the results of the cross-language validation of machine learning models. The robustness of the features and hypotheses arising from the classification results need to be verified in a follow-up study where data will be homogeneous and subjects preferably of the same age and level of PD progression.

## 5 Conclusion

We analyzed in detail the behaviour of acoustic speech features in different languages. The aim was to explore digital speech biomarkers of PD and determine which are independent of the speaker’s language yet have high discrimination power.

Our statistical analysis found that approximately one-third of the significant features did not meet the conditions for language robustness. The most successful biomarkers in this sense include lower prominence of the second formant, monopitch, and a lower number of pauses detected during text reading. These biomarkers also performed best during classification using machine learning, both in the single language models and the model using all languages’ features together. Classification accuracies ranged from 67 % to 85 %, depending on the language. The model trained with features of all languages together achieved a sensitivity of 73 % and a specificity of 76 %.

This work contributes insights into the field of objective assessment of speech affected by hypokinetic dysarthria and automated diagnosis of PD. It is the first study to use the concept of language robustness, and through the detailed exploration of speech features using both statistics and machine learning, it proposes several digital speech biomarkers that have the potential to be language-independent and that could be possibly used in eHealth/mHealth applications.

## Data Availability

Data are protected by privacy and security law. Code with used algorithms can be found in the GitHub repository [Multilingual speech analysis] [https://github.com/BDALab/multilingual_speechanalysis].

## Acknowledgement

This work was supported by the Czech Ministry of Health under grant no. NU20-04-00294, by EU – Next Generation EU (project no. LX22NPO5107 (MEYS)), and by the European Union’s Horizon 2020 research and innovation program under the Marie Sk-lodowska-Curie grant agreement no. 734718 (CoBeN).

## Authorship contribution

**Daniel Kovac**: conceptualization, data processing, acoustic features extraction, machine learning, GitHub repository management, article writing. **Jiri Mekyska**: conceptualization, acoustic features extraction, statistical analysis, machine learning, article writing. **Vered Aharonson**: conceptualization, data provision. **Pavol Harar**: statistical analysis, machine learning, GitHub repository management. **Zoltan Galaz**: machine learning. **Steven Rapcsak**: data provision. **Juan Rafael Orozco-Arroyave**: data provision. **Lubos Brabenec**: data provision. **Irena Rektorova**: conceptualization, critical reading of the manuscript draft.

## Notes

### Competing Interest Statement

The authors have declared no competing interest.

### Author Declarations

The Research Ethics Committee (REC) of Masaryk University chaired by prof. RNDr. Renata Veselska, Ph.D., M.Sc. gave ethical approval for this work.

### Summary of Updates

Partial Spearman's correlations with 'language' set as a covariate to evaluate relationships between speech and clinical data (Table 5).

## References

[1] N. Muñoz-Vigueras, E. Prados-Román, M. C. Valenza, M. Granados-Santiago, I. Cabrera-Martos, J. Rodríguez-Torres, and I. Torres-Sánchez, “Speech and language therapy treatment on hypokinetic dysarthria in parkinson disease: Systematic review and meta-analysis,” Clinical Rehabilitation, vol. 35, no. 5, pp. 639–655, 2021.

[2] A. Rohl, S. Gutierrez, K. Johari, J. Greenlee, K. Tjaden, and A. Roberts, “Chapter 7 - speech dysfunction, cognition, and parkinson’s disease,” in Cognition in Parkinson’s Disease (N. S. Narayanan and R. L. Albin, eds.), vol. 269 of Progress in Brain Research, pp. 153–173, Elsevier, 2022.

[3] T. Tykalová, J. Rusz, J. Švihlík, S. Bancone, A. Spezia, and M. T. Pellecchia, “Speech disorder and vocal tremor in postural instability/gait difficulty and tremor dominant subtypes of parkinson’s disease,” Journal of Neural Transmission, vol. 127, no. 9, pp. 1295–1304, 2020.

[4] M. Cernak, J. R. Orozco-Arroyave, F. Rudzicz, H. Christensen, J. C. Vásquez-Correa, and E. Nöth, “Characterisation of voice quality of parkinson’s disease using differential phonological posterior features,” Computer Speech & Language, vol. 46, pp. 196–208, 2017.

[5] Z. Thijs and C. R. Watts, “Perceptual characterization of voice quality in nonadvanced stages of parkinson’s disease,” Journal of Voice, 2020.

[6] R. B. Hoodin and H. R. Gilbert, “Nasal airflows in parkinsonian speakers,” Journal of Communication Disorders, vol. 22, no. 3, pp. 169–180, 1989.

[7] A. M. Goberman, M. Blomgren, and E. Metzger, “Characteristics of speech disfluency in parkinson disease,” Journal of Neurolinguistics, vol. 23, no. 5, pp. 470–478, 2010.

[8] F. S. Juste, F. C. Sassi, J. B. Costa, and C. R. F. de Andrade, “Frequency of speech disruptions in parkinson’s disease and developmental stuttering: A comparison among speech tasks,” Plos one, vol. 13, no. 6, p. e0199054, 2018.

[9] V. L. Hammen and K. M. Yorkston, “Speech and pause characteristics following speech rate reduction in hypokinetic dysarthria,” Journal of communication disorders, vol. 29, no. 6, pp. 429–445, 1996.

[10] K. Tjaden and G. Wilding, “Effects of speaking task on intelligibility in parkinson’s disease,” Clinical Linguistics & Phonetics, vol. 25, no. 2, pp. 155–168, 2011.

[11] S. G. Adams, A. Dykstra, M. Jenkins, and M. Jog, “Speech-to-noise levels and conversational intelligibility in hypophonia and parkinson’s disease,” Journal of medical speech-language pathology, vol. 16, no. 4, pp. 165–173, 2008.

[12] F. L. Darley, A. E. Aronson, and J. R. Brown, “Differential diagnostic patterns of dysarthria,” Journal of speech and hearing research, vol. 12, no. 2, pp. 246–269, 1969.

[13] J. R. Duffy, Motor speech disorders e-book: Substrates, differential diagnosis, and management. Elsevier Health Sciences, 2019.

[14] O. Hornykiewicz, “Biochemical aspects of Parkinson’s disease,” Neurology, vol. 51, no. 2 Suppl 2, pp. S2–S9, 1998.

[15] K. Ho, R. Iansek, C. Marigliani, J. L. Bradshaw, and S. Gates, “Speech impairment in a large sample of patients with parkinson’s disease,” Behavioural neurology, vol. 11, no. 3, pp. 131–137, 1998.

[16] O.-B. Tysnes and A. Storstein, “Epidemiology of parkinson’s disease,” Journal of neural transmission, vol. 124, pp. 901–905, 2017.

[17] G. DeMaagd and A. Philip, “Parkinson’s disease and its management: part 1: disease entity, risk factors, pathophysiology, clinical presentation, and diagnosis,” Pharmacy and therapeutics, vol. 40, no. 8, p. 504, 2015.

[18] J. Parkinson, “An essay on the shaky palsy,” London: Sherwood, Neely and Jones, pp. 1–6, 1817.

[19] C. McDonald, G. Gordon, A. Hand, R. W. Walker, and J. M. Fisher, “200 years of parkinson’s disease: what have we learnt from james parkinson?,” Age and ageing, vol. 47, no. 2, pp. 209–214, 2018.

[20] W. Poewe, “Global scales to stage disability in pd: the hoehn and yahr scale,” Rating Scales Parkinsons Dis, pp. 115–122, 2012.

[21] T. L. Whitehill, “Studies of chinese speakers with dysarthria: informing theoretical models,” Folia Phoniatrica et Logopaedica, vol. 62, no. 3, pp. 92–96, 2010.

[22] H. Hazan, D. Hilu, L. Manevitz, L. O. Ramig, and S. Sapir, “Early diagnosis of Parkinson’s disease via machine learning on speech data,” in 2012 IEEE 27th Convention of Electrical and Electronics Engineers in Israel, pp. 1–4, IEEE, 2012.

[23] J. Orozco-Arroyave, F. Hönig, J. Arias-Londoño, J. Vargas-Bonilla, K. Daqrouq, S. Skodda, J. Rusz, and E. Nöth, “Automatic detection of Parkinson’s disease in running speech spoken in three different languages,” The Journal of the Acoustical Society of America, vol. 139, no. 1, pp. 481–500, 2016.

[24] Y. Kim and Y. Choi, “A cross-language study of acoustic predictors of speech intelligibility in individuals with Parkinson’s disease,” Journal of Speech, Language, and Hearing Research, vol. 60, no. 9, pp. 2506–2518, 2017.

[25] L. Moro-Velazquez, J. A. Gomez-Garcia, J. I. Godino-Llorente, F. Grandas-Perez, S. Shattuck-Hufnagel, V. Yagüe-Jimenez, and N. Dehak, “Phonetic relevance and phonemic grouping of speech in the automatic detection of Parkinson’s disease,” Scientific reports, vol. 9, no. 1, pp. 1–16, 2019.

[26] J. C. Vásquez-Correa, T. Arias-Vergara, C. D. Rios-Urrego, M. Schuster, J. Rusz, J. R. Orozco-Arroyave, and E. Nöth, “Convolutional neural networks and a transfer learning strategy to classify Parkinson’s disease from speech in three different languages,” in Iberoamerican Congress on Pattern Recognition, pp. 697–706, Springer, 2019.

[27] J. Rusz, J. Hlavnička, M. Novotný, T. Tykalová, A. Pelletier, J. Montplaisir, J.-F. Gagnon, P. Dušek, A. Galbiati, S. Marelli, et al., “Speech biomarkers in rapid eye movement sleep behavior disorder and parkinson disease,” Annals of neurology, vol. 90, no. 1, pp. 62–75, 2021.

[28] A. S. Ozbolt, L. Moro-Velazquez, I. Lina, A. A. Butala, and N. Dehak, “Things to consider when automatically detecting parkinson’s disease using the phonation of sustained vowels: Analysis of methodological issues,” Applied Sciences, vol. 12, no. 3, p. 991, 2022.

[29] L. Brabenec, P. Klobusiakova, P. Simko, M. Kostalova, J. Mekyska, and I. Rektorova, “Non-invasive brain stimulation for speech in parkinson’s disease: A randomized controlled trial,” Brain Stimulation, vol. 14, no. 3, pp. 571–578, 2021.

[30] Z. Galaz, J. Mekyska, Z. Mzourek, Z. Smekal, I. Rektorova, I. Eliasova, M. Kostalova, M. Mrackova, and D. Berankova, “Prosodic analysis of neutral, stress-modified and rhymed speech in patients with parkinson’s disease,” Computer methods and programs in biomedicine, vol. 127, pp. 301–317, 2016.

[31] D. Kovac, J. Mekyska, Z. Galaz, L. Brabenec, M. Kostalova, S. Z. Rapcsak, and I. Rektorova, “Multilingual analysis of speech and voice disorders in patients with parkinson’s disease,” in 2021 44th International Conference on Telecommunications and Signal Processing (TSP), pp. 273–277, 2021.

[32] J. R. Orozco-Arroyave, J. D. Arias-Londoño, J. F. Vargas-Bonilla, M. C. González-Rátiva, and E. Nöth, “New Spanish speech corpus database for the analysis of people suffering from Parkinson’s disease,” in Proceedings of the Ninth International Conference on Language Resources and Evaluation (LREC’14), (Reykjavik, Iceland), pp. 342–347, European Language Resources Association (ELRA), May 2014.

[33] G. Dimauro and F. Girardi, “Italian parkinson’s voice and speech,” 2019.

[34] L. Brabenec, J. Mekyska, Z. Galaz, and I. Rektorova, “Speech disorders in parkinson’s disease: early diagnostics and effects of medication and brain stimulation,” Journal of neural transmission, vol. 124, no. 3, pp. 303–334, 2017.

[35] L. Moro-Velazquez, J. A. Gomez-Garcia, J. D. Arias-Londoño, N. Dehak, and J. I. Godino-Llorente, “Advances in parkinson’s disease detection and assessment using voice and speech: A review of the articulatory and phonatory aspects,” Biomedical Signal Processing and Control, vol. 66, p. 102418, 2021.

[36] L. Moro-Velazquez and N. Dehak, “A review of the use of prosodic aspects of speech for the automatic detection and assessment of parkinson’s disease,” in Automatic Assessment of Parkinsonian Speech Workshop, pp. 42–59, Springer, 2019.

[37] P. Corcoran, A. Hensman, and B. Kirkpatrick, “Glottal flow analysis in parkinsonian speech.,” in BIOSIGNALS, pp. 116–123, 2019.

[38] M. T. Todd, L. E. Nystrom, and J. D. Cohen, “Confounds in multivariate pattern analysis: theory and rule representation case study,” Neuroimage, vol. 77, pp. 157–165, 2013.

[39] K. N. Stevens, S. Y. Manuel, S. Shattuck-Hufnagel, and S. Liu, “Implementation of a model for lexical access based on features.,” in ICSLP, no. October, pp. 499–502, 1992.

[40] J. Slifka, K. N. Stevens, S. Manuel, and S. Shattuck-Hufnagel, “A landmark-based model of speech perception: History and recent developments,” From Sound to Sense, pp. 85–90, 2004.

[41] S. Boyce, H. Fell, and J. MacAuslan, “Speechmark: Landmark detection tool for speech analysis,” in Thirteenth Annual Conference of the International Speech Communication Association, 2012.

[42] J. Rusz, T. Tykalova, M. Novotny, D. Zogala, K. Sonka, E. Ruzicka, and P. Dusek, “Defining speech subtypes in de novo parkinson disease: response to long-term levodopa therapy,” Neurology, vol. 97, no. 21, pp. e2124–e2135, 2021.

[43] J. Rusz and T. Tykalova, “Does cognitive impairment influence motor speech performance in de novo parkinson’s disease?,” Movement Disorders, vol. 36, no. 12, pp. 2980–2982, 2021.

[44] M. García, T. Arias-Vergara, J. C Vasquez-Correa, E. Nöth, M. Schuster, A. E. Welch, Y. Bocanegra, A. Baena, and J. R. Orozco-Arroyave, “Cognitive determinants of dysarthria in parkinson’s disease: an automated machine learning approach,” Movement Disorders, vol. 36, no. 12, pp. 2862–2873, 2021.

[45] I. Rektorova, J. Mekyska, E. Janousova, M. Kostalova, I. Eliasova, M. Mrackova, D. Berankova, T. Necasova, Z. Smekal, and R. Marecek, “Speech prosody impairment predicts cognitive decline in parkinson’s disease,” Parkinsonism & related disorders, vol. 29, pp. 90–95, 2016.

[46] J. Rusz, T. Tykalovy, M. Novotny, D. Zogala, E. Ruzicka, and P. Dusek, “Automated speech analysis in early untreated parkinson’s disease: relation to gender and dopaminergic transporter imaging,” European Journal of Neurology, vol. 29, no. 1, pp. 81–90, 2022.

